# How influenza vaccination changed over the COVID-19 pandemic

**DOI:** 10.1101/2023.03.21.23287546

**Authors:** Yong Yang

## Abstract

**Background:** Vaccination for seasonal influenzas is particularly important during the COVID-19 pandemic, but the influenza vaccination coverage in the U.S. was far lower than the targeted rate.

**Objective:** To examine how people’s actual uptake of the influenza vaccine and the disparity of the vaccination changed during the pandemic.

**Methods:** A survey was conducted online in November 2022. Respondents were asked for influenza vaccination during each of the three latest seasons, prior influenza vaccination history, and COVID-19 vaccination. A linear regression model was used to estimate how the respondents’ change in influenza vaccination was associated with their demographics, COVID-19 vaccination status, and other related variables.

**Results:** Nearly 70% of US adults had influenza vaccine each season during past the three seasons of the COVID-19 pandemic. The prevalence of influenza vaccination varied markedly across demographics. Non-Hispanic Black, Hispanic, and people with low educational attainment were more likely to see relatively negative changes in their level of influenza vaccination. Respondents who uptook their COVID-19 vaccine in 2022 increased their level of influenza vaccine more than those who uptook the vaccine in 2021.

**Conclusions:** Our study indicated that influenza vaccination increased during the pandemic compared with before the pandemic. The disparity of influenza vaccination by race/ethnicity and socioeconomic status may enlarge during the pandemic. Tailored interventions were needed to target some groups to promote their vaccination uptake.

## Introduction

Influenza is a highly contagious respiratory disease that spreads worldwide in annual outbreaks, causing significant health burdens including morbidity and mortality [1, 2]. Influenza vaccination every year is the best way to protect individuals from Influenza [1-4]. During the ongoing COVID-19 pandemic, vaccination for seasonal influenzas is particularly important because both diseases peak in Winter, and they may spread simultaneously, thus the risk of co-infection and twindemic may pose a serious burden on healthcare systems [5-7].

Recent data indicated that influenza vaccination coverage may slightly increase in the US during the COVID-19 pandemic. The influenza vaccination coverages among adults ≥18 years were 50.2% and 49.4% in the seasons of 2020-21 and 2021-22, respectively [8], compared with a fluctuation of a range of 37.1-45.3% between 2009 and 2018, with a peak of 48.4% in the season of 2019-20 [8]. However, these rates were far lower than the targeted rate of 70% according to the Healthy People 2030 plan [9]. Further, significant disparities remain in influenza vaccination, as the rates are is lower among Hispanics and non-Hispanic Blacks compared with non-Hispanic Whites, and are lower among groups with lower socioeconomic status compared with their counterparts [10, 11].

Influenza vaccination may change during the pandemic. Initial evidence showed that influenza vaccination may reduce the risk of COVID-19 infection and its severity [17-19]. Evidence showed that people with a history of influenza vaccinations were more likely to uptake the COVID-19 vaccine and the demographic correlates of vaccinations were roughly similar between influenza and COVID-19 [12-14]. At the same time, It is possible that factors associated with COVID-19 vaccination that emerged during the pandemic from both positive (e.g., the effectiveness of COVID-19 to protect from COVID-19 infection and severity) and negative direction (e.g., safety concerns and mistrust of COVID-19 vaccines or government) may change people’ intention to vaccination in general and thus affect influenza vaccination afterward [15]. One meta-analysis study [16] found that the intention to vaccinate against influenza has increased during the COVID-19 pandemic globally.

Our understanding of how people’s actual uptake of influenza vaccine changed during the pandemic, particularly, how the change may vary with the population and how the uptake of COVID-19 vaccine may impact people’s uptake of influenza vaccine is limited. This study aimed to address the above knowledge gap.

## Methods

A survey was conducted online using a crowdsourcing platform in November 2022 and adults who were 18 years old and above and currently lived in the US were eligible to attend the survey. The Institutional Review Board at the University of Memphis approved this study. Respondents were asked for their influenza vaccination during each of the three latest seasons using three questions as follows: “Did you get a flu vaccine between Fall 2020 and Spring 2021?”, “Did you get a flu vaccine between Fall 2021 and Spring 2022?” and “Did you get a flu vaccine or plan to have it for this flu season, that is, between Fall 2022 to Spring 2023?”. Respondents were asked for their prior influenza vaccination history using the question “prior to the COVID-19 pandemic, how often did you get a flu vaccine?”, with four levels including every year or almost every year, some years but not others, only once, and never. Respondents were also asked if they had been infected by COVID-19 and their COVID-19 vaccination status using the question “Have you got COVID-19 vaccine? either the first shot or both shots?” Those answered yes will be asked for the time of their vaccination with four options including (1) the first half of 2021; (2) the second half of 2021; (3) the first half of 2022; and (4) the second half of 2022. Besides demographics, respondents were asked for their insurance status and political views (either Democratic, Republican, Independent, or others).

As data analysis, first, we estimated the percentage of people who uptook the influenza vaccine each season during the pandemic, that is, from 2020 to 2023, by adding weights to ensure the demographics of the weighted sample match with the US general population according to the 2020 Census. Second, we analyzed the change in influenza vaccination during the pandemic compared with the prior influenza vaccination history before the pandemic. The total number of influenza vaccination during the three influenza seasons between 2020 and 2023 ranged between 0 (i.e., did not uptake influenza vaccine in any of the three seasons) and 3 (i.e., uptook influenza vaccine in all three seasons). Correspondingly, the four levels of prior influenza vaccination history were coded as: 0 for never, 1 for only once, 2 for some years but not others, and 3 for every year or almost every year. These codes and numbers were roughly consistent and comparable to reflect the degree of respondents’ influenza vaccination before and during the pandemic. Thus, we used their difference as a variable to present the change in influenza vaccination over the pandemic, with a positive value indicating an increased uptake. A linear regression model was used to estimate how the respondents’ change in influenza vaccination was associated with their demographics, COVID-19 vaccination status, and other related variables.

## Results

The sample included 1399 participants, with males, younger adults, Hispanic, and those with high educational attainment overrepresented (see Table 1). There were 69.8% of the respondents had been infected by COVID-19, 85.4% had COVID-19 vaccination in 2021, and 12% had COVID-19 vaccination in 2022. With weights, the mean prevalence of influenza vaccination was 69.5% during the three seasons between 2020 and 2023, with 72.0%, 67.0%, and 69.0% for each season, respectively. As Table 1 shows, the prevalence of influenza vaccination varied markedly across demographics, with some groups lower than 50% such as those with household income lower than $24,999 per year (48.1%), Blacks (44.9%), those who never uptook influenza vaccine prior to the pandemic (12.9%), and those who did not uptake COVID-19 vaccine (2.0%).

**Table 1.** Characteristics of respondents (N=1399) and the averaged weighted prevalence of influenza vaccination for three seasons from 2020 to 2023.

| Category | Item | Percent (%) | The prevalence of flu vaccination |
| --- | --- | --- | --- |
| All |  |  | 69.5 |
| Gender | Male | 71.7 | 75.9 |
|  | Female | 28.3 | 63.2 |
| Age | 18-34 | 67.1 | 63.5 |
|  | 35-59 | 28.7 | 66.4 |
|  | 60 and above | 4.3 | 78.6 |
| Race /ethnicity | White | 63.7 | 71.2 |
|  | Black | 4.2 | 44.9 |
|  | Asian | 6.3 | 72.1 |
|  | Hispanic | 22.7 | 80.3 |
|  | Others | 3.1 | 74.4 |
| Educational attainment | High school and below | 9.4 | 62.2 |
|  | Above high school and below bachelor | 4.4 | 64.4 |
|  | Bachelor and above | 86.2 | 80.2 |
| Household Income | Less than \$24,999 | 11.6 | 48.1 |
| | \$25,000 to \$49,999 | 28.4 | 70.8 |
| | \$50,000 to \$74,999 | 36.1 | 86.7 |
| | \$75,000 to \$99,999 | 17.8 | 79.0 |
| | \$100,000 or more | 6.2 | 67.4 |
| Political view | Democrat | 48.7 | 73.0 |
|  | Independent | 15.7 | 62.8 |
|  | Republican | 35.7 | 69.0 |
| With health insurance | No | 18.9 | 65.7 |
|  | Yes | 81.1 | 69.8 |
| If infected by COVID-19 | Yes | 69.8 | 77.0 |
|  | No | 30.2 | 56.2 |
| The time of taking COVID-19 vaccine | No | 2.6 | 2.0 |
|  | The first half of 2021 | 46.0 | 66.6 |
|  | The second half of 2021 | 39.4 | 78.7 |
|  | The first half of 2022 | 9.6 | 86.2 |
|  | The second half of 2022 | 2.4 | 81.3 |
| Prior influenza vaccination | Every year or almost every year | 32.3 | 85.7 |
|  | Some years | 33.0 | 79.0 |
|  | Only once | 29.6 | 59.4 |
|  | Never | 5.2 | 12.9 |
| Number of influenza vaccination between 2020 and 2022 | 3 | 58.8 |  |
|  | 2 | 23.7 |  |
|  | 1 | 9.8 |  |
|  | 0 | 7.8 |  |

Note: the weighted prevalence of flu vaccination for 72.0% for the season of 2020-2021, 67.0% for the season of 2021-2022, and 69.0% for the season of 2022-2023

Table 2 shows that the level of influenza vaccination decreased among females and all minorities except non-Hispanic Asian compared with their counterparts. People with a higher level of educational attainment, those with health insurance, and who were infected by COVID-19 increased their level of influenza vaccination compared with those with a lower level of educational attainment, those without health insurance, and those who have not been infected by COVID-19, respectively. Compared with respondents with a Democratic political view, those with a Republican view decreased their level of influenza vaccination.

**Table 2.** Result of a linear regression model for the change in influenza vaccination level before the COVID-19 pandemic compared with the three seasons from 2020 to 2023.

| Variables | Items | <sup>1</sup>Change in influenza vaccination level |
| --- | --- | --- |
| Gender | Male (ref) | 0 |
|  | Female | -0.26(-0.37,-0.15)** |
| Age groups | 18-34 (ref) | 0 |
|  | 35-59 | 0.03(-0.09,0.15) |
|  | 60 and above | 0.14(-0.01,0.28) |
| Race /ethnicity | Non-Hispanic White (ref) | 0 |
|  | Non-Hispanic Black | -0.64(-0.79,-0.49)** |
|  | Non-Hispanic Asian | -0.05(-0.28,0.17) |
|  | Hispanic | -0.25(-0.39,-0.11)** |
|  | Others and mixed | -0.41(-0.77,-0.05)* |
| <sup>2</sup> Education attainment |  | 0.17(0.1,0.24)** |
| <sup>3</sup> Household income |  | -0.09(-0.12,-0.05)** |
| Political view | Democrat (ref) | 0 |
|  | Independent and others | -0.03(-0.17,0.11) |
|  | Republican | -0.15(-0.27,-0.04)* |
| With health insurance | No (ref) | 0 |
|  | Yes | 0.17(0.03,0.3)* |
| Prior influenza vaccination | Every year or almost every year (ref) | 0 |
|  | Some years | 0.71(0.58,0.84)** |
|  | Only once | 1.35(1.22,1.49)** |
|  | Never | 1.1(0.93,1.27)** |
| If infected by COVID-19 | No (ref) | 0 |
|  | Yes | 0.41(0.30,0.52)** |
| The time of taking COVID-19 vaccine | No (ref) | 0 |
|  | The first half of 2021 | 0.93(0.68,1.19)** |
|  | The second half of 2021 | 1.15(0.88,1.41)** |
|  | The first half of 2022 | 1.25(0.94,1.55)** |
|  | The second half of 2022 | 1.20(0.62,1.78)** |

Notes: boldface indicates statistical significance, with \* for $p < 0.05$ , and \*\* for $p < 0.01$
<sup>1</sup>The total number of influenza vaccination during the three influenza seasons between 2020 and 2023 ranged between 0 (i.e., did not uptake influenza vaccine in any of the three seasons) and 3 (i.e., uptook influenza vaccine in all three seasons). Correspondingly, the four levels of prior influenza vaccination history were coded as: 0 for never, 1 for only once, 2 for some years but not others, and 3 for every year or almost every year. These codes and numbers were roughly consistent and comparable to reflect the degree of respondents' influenza vaccination before and during the pandemic. Thus, we used their difference as a variable to present the change in influenza vaccination over the pandemic, with a positive value indicating an increased uptake.
<sup>2</sup>Educational attainment was categorized into three levels: 1. high school or less; 2. less than bachelor and more than high school; and 3. bachelor or more.
<sup>3</sup>Household income was categorized into five levels: 1. Less than \$24,999; 2. \$25,000 to \$49,999; 3. \$50,000 to \$74,999; 4. \$75,000 to \$99,999; and 5. \$100,000 or more.

People with a low level of prior influenza vaccination history were more likely to observe a relative increase compared with their counterparts. However, those who uptook the influenza vaccine once before increased more than those who never had the vaccine before. Compared with those who did not have the COVID-19 vaccine during the pandemic, responses who had the COVID-19 vaccine significantly increased their level of influenza vaccine but those who uptook their COVID-19 vaccine later (i.e., in 2022) increased more than those who uptook the vaccine earlier (i.e., in 2021).

## Discussions

Our study showed nearly 70% of US adults had influenza vaccine during past the several seasons of the COVID-19 pandemic. Our prevalence may be overestimated because our participants were not representative of the U.S. population. Our participants were highly educated and were recruited from the Internet thus the bias may not be eliminated fully through the weighting (e.g., those who had no access to the Internet could not be presented). However, even considering the possible bias, the prevalence was significantly higher than it was before the pandemic and we observed high prevalence among certain groups, for example, 59.4% among those who only had influenza vaccine once before the pandemic. This indicated the influenza vaccine increased during the pandemic compared with before the pandemic.

Our results showed the change in influenza vaccination was not evenly distributed, and some groups including Non-Hispanic Black, Hispanic, and people with low educational attainment, who were less likely to uptake influenza vaccine before the pandemic were more likely to see relatively negative changes in their level of influenza vaccination. This indicated that the disparity of influenza vaccination by race/ethnicity and socioeconomic status may enlarge over the pandemic. The negative change of influenza vaccination among Republicans, compared with Democrats, is consistent with a recent study that confirmed the decline of intentions of uptaking influenza vaccine and the general attitudes towards vaccine among Republicans [20].

It was not surprising to see that people who uptook the influenza vaccine every year or almost every year before the pandemic were the group that had the least relatively positive change compared with other groups because there was no room for this group to improve (they already had the highest level). But it was interesting to see that it was the group who uptook the influenza vaccine once before the pandemic, rather than the group who never had the influenza vaccine before (despite their lowest level of prior history and thus the large potential compared with other groups), had the largest increase of the level of influenza vaccine. This indicated that the group who never had influenza vaccine before the pandemic, either because they were against the vaccine or could not uptake it due to other reasons (e.g., physical conditions), was also the most unlikely to change. Another interesting finding was that the respondents who uptook their COVID-19 vaccine in 2022 increased their level of influenza vaccine more than those who uptook the vaccine in 2021. A possible explanation is that those who were reluctant to uptake the COVID-19 vaccine initially but eventually uptook the vaccine may increase their uptake of the influenza vaccine as a measure to keep them away from COVID-19 infection or this group after they finally switch to uptake the COVID-19 vaccine from the initial reluctance, also increased their uptake of other vaccines, this is consistent with the finding of another study that individuals who did not uptake influenza vaccine often before the pandemic, when they received COVID-19 vaccine, they were more likely to increase their the influenza vaccine [14].

Overall our study showed the prevalence of influenza vaccination may increase during the pandemic compared with before but the disparity of influenza vaccination in the population may enlarge as the groups such as minorities and low SES who were typically less likely to uptake influenza vaccine were also more likely to decrease their level of vaccination compared with their counterparts. The groups including Republicans, those who never uptake the influenza vaccine before the pandemic, and those who did not take the COVID-19 vaccine were the group that was the most unlikely to increase their uptake of the influenza vaccine. Tailored intervention programs were needed to specifically target these groups to promote their vaccination uptake, and this is crucial to promote influenza vaccination coverage overall and also decrease the disparity among the population.

## Data Availability

All data produced in the present study are available upon reasonable request to the authors

## References

1. CDC. Disease Burden of Flu. 2022 [cited 2022 Dec 29, 2022]; Available from: https://www.cdc.gov/flu/about/burden/index.html.

2. WHO. Fact sheet of Influenza (Seasonal). 2018 [cited 2022 Dec 29]; Available from: https://www.who.int/news-room/fact-sheets/detail/influenza-(seasonal).

3. Uyeki, T.M., Influenza. Annals of Internal Medicine, 2017. 167(5): p. ITC33–ITC48.

4. Becker, T., H. Elbahesh, L.A. Reperant, G.F. Rimmelzwaan, and A.D.M.E. Osterhaus, Influenza Vaccines: Successes and Continuing Challenges. The Journal of Infectious Diseases, 2021. 224(Supplement_4): p. S405–S419.

5. Wang, R., L. Tao, N. Han, J. Liu, C. Yuan, L. Deng, C. Han, F. Sun, L. Chi, M. Liu, and J. Liu, Acceptance of seasonal influenza vaccination and associated factors among pregnant women in the context of COVID-19 pandemic in China: a multi-center cross-sectional study based on health belief model. BMC Pregnancy Childbirth, 2021. 21(1): p. 745.

6. Rubin, R., The Dreaded “Twindemic” of Influenza and COVID-19 Has Not Yet Materialized— Might This Be the Year? JAMA, 2022. 328(15): p. 1488–1489.

7. Dadashi, M., S. Khaleghnejad, P. Abedi Elkhichi, M. Goudarzi, H. Goudarzi, A. Taghavi, M. Vaezjalali, and B. Hajikhani, COVID-19 and Influenza Co-infection: A Systematic Review and Meta-Analysis. Frontiers in Medicine, 2021. 8.

8. CDC. Influenza Coverage by Season. 2022; Available from: https://www.cdc.gov/flu/fluvaxview/coverage-by-season.htm.

9. U.S. Department of Health and Human Services. Healthy People 2030. 2022 [cited 2022 Dec 29, 2022]; Available from: https://health.gov/healthypeople.

10. Lu, P.-j., M.-C. Hung, A.C. O’Halloran, H. Ding, A. Srivastav, W.W. Williams, and J.A. Singleton, Seasonal Influenza Vaccination Coverage Trends Among Adult Populations, U.S., 2010–2016. American Journal of Preventive Medicine, 2019. 57(4): p. 458–469.

11. Black, C.L., A. O’Halloran, and M.-C. Hung, Vital Signs: Influenza Hospitalizations and Vaccination Coverage by Race and Ethnicity—United States, 2009–10 Through 2021–22 Influenza Seasons. Morb Mortal Wkly Rep, 2022. 71: p. 1366–1373.

12. Fisher, K.A., S.J. Bloomstone, J. Walder, S. Crawford, H. Fouayzi, and K.M. Mazor, Attitudes Toward a Potential SARS-CoV-2 Vaccine : A Survey of U.S. Adults. Annals of internal medicine, 2020. 173(12): p. 964–973.

13. Yang, Y., A. Dobalian, and K.D. Ward, COVID-19 Vaccine Hesitancy and Its Determinants Among Adults with a History of Tobacco or Marijuana Use. Journal of Community Health, 2021.

14. Parker, A.M., S. Atshan, M.M. Walsh, C.A. Gidengil, and R. Vardavas, Association of COVID-19 Vaccination With Influenza Vaccine History and Changes in Influenza Vaccination. JAMA Network Open, 2022. 5(11): p. e2241888–e2241888.

15. Leuchter, R.K., N.J. Jackson, J.N. Mafi, and C.A. Sarkisian, Association between Covid-19 Vaccination and Influenza Vaccination Rates. New England Journal of Medicine, 2022. 386(26): p. 2531–2532.

16. Kong, G., N.-A. Lim, Y.H. Chin, Y.P.M. Ng, and Z. Amin, Effect of COVID-19 Pandemic on Influenza Vaccination Intention: A Meta-Analysis and Systematic Review. Vaccines, 2022. 10(4): p. 606.

17. Tayar, E., S. Abdeen, M. Abed Alah, H. Chemaitelly, I. Bougmiza, H.H. Ayoub, A.H. Kaleeckal, A.N. Latif, R.M. Shaik, H.E. Al-Romaihi, M.H. Al-Thani, R. Bertollini, L.J. Abu-Raddad, and A. Al-Khal, Effectiveness of influenza vaccination against SARS-CoV-2 infection among healthcare workers in Qatar. J Infect Public Health, 2023. 16(2): p. 250–256.

18. Hosseini-Moghaddam, S.M., S. He, A. Calzavara, M.A. Campitelli, and J.C. Kwong, Association of Influenza Vaccination With SARS-CoV-2 Infection and Associated Hospitalization and Mortality Among Patients Aged 66 Years or Older. JAMA Network Open, 2022. 5(9): p. e2233730–e2233730.

19. Conlon, A., C. Ashur, L. Washer, K.A. Eagle, and M.A. Hofmann Bowman, Impact of the influenza vaccine on COVID-19 infection rates and severity. American Journal of Infection Control, 2021. 49(6): p. 694–700.

20. Fridman, A., R. Gershon, and A. Gneezy, COVID-19 and vaccine hesitancy: A longitudinal study. PLOS ONE, 2021. 16(4): p. e0250123.

